# Submaximal Exercise Testing to Dose High-Intensity Interval Training After Stroke: The FAST Randomized Clinical Trial

**DOI:** 10.64898/2026.03.17.26348646

**Authors:** Bria L. Bartsch, Amanda Engler, Noah Schneider, Amanda J. Britton-Carpenter, Tyler Baldridge, Robert N. Montgomery, Eric D. Vidoni, Alexandra Moores, Elyse S. Vetter, Emily M. Hazen, Michael Abraham, Sandra A. Billinger

**Author notes:** <u>Corresponding Author:</u> S Billinger, PhD, 3901 Rainbow Blvd, Mail Stop 3051, Kansas City, KS, 66160.

## Abstract

**Importance:** High-intensity interval training (HIIT) improves peak oxygen uptake (VO*_2_*peak) and walking post-stroke. However, previous HIIT trials have primarily implemented maximal exercise testing, limiting clinical implementation.

**Objective:** Evaluate the preliminary efficacy of HIIT, compared to moderate-intensity continuous training (MICT) using a submaximal exercise test. Hypothesis: HIIT will produce greater improvements than MICT in VO*_2_*peak, vascular measures, and walking outcomes.

**Design:** This was a randomized preliminary efficacy trial conducted between July 2023 and December 2025.

**Setting:** University of Kansas Medical Center.

**Participants:** Participants with chronic stroke, 20-85 years of age, were randomized to HIIT or MICT.

**Intervention:** HIIT and MICT were performed on a total-body recumbent stepper 3 times per week for 4 weeks, with intensity prescribed using peak power output (PPO) to achieve target heart rate zones derived from a submaximal exercise test. HIIT was performed for 25 minutes with 1-minute vigorous-intensity intervals (65-95% PPO) interspersed with 1-minute active recovery intervals. MICT was performed continuously at 45-65% PPO for 25 minutes.

**Main Outcomes:** The primary outcome was change in predicted VO*_2_*peak. Secondary outcomes included middle cerebral artery velocity, peripheral vascular function, and arterial stiffness with gait speed and walking endurance as tertiary outcomes.

**Results:** Forty-nine participants (HIIT: n=25, MICT: n=24) were randomized (62.4(12.5) years, 42.9% female), attended 99.5(2.0)% of sessions, and achieved target intensity zones. No study-related serious adverse events occurred. Our results showed no significant between-group differences (p=0.54) for study outcomes. Both groups significantly improved VO*_2_*peak (HIIT: +1.13 mL•kg^-1^•min^-1^ (95% CI: 0.05-2.21), p=0.04; MICT: +1.58 mL•kg^-1^•min^-1^ (95% CI: 0.18-2.97), p=0.03) and with fast gait speed and walking endurance. Peripheral vascular function significantly improved following HIIT.

**Conclusions and Relevance:** HIIT can be safely implemented in individuals with chronic stroke using a submaximal exercise test. Both HIIT and MICT elicited clinically meaningful gains in VO_2_peak and walking. However, only HIIT led to a significant improvement in peripheral vascular function, suggesting a biologic signal for intensity-dependent vascular adaptation.

**Trial Registration:** ClinicalTrials.gov identifier: NCT05936008.

**Key Points:** *Question:* In individuals with chronic stroke, does high-intensity interval training (HIIT) improve predicted VO₂peak more than moderate-intensity continuous training (MICT)?

*Findings:* In this randomized clinical trial of 49 participants with chronic stroke, both HIIT and MICT achieved prescribed intensity targets with high adherence and resulted in clinically meaningful improvements in predicted VO₂peak and walking outcomes after 4 weeks, with no significant between-group difference in our primary outcome of VO₂peak.

*Meaning:* These findings suggest that when aerobic exercise is prescribed to achieve target intensity, both HIIT and MICT produce meaningful improvements in fitness and walking after stroke, supporting the importance of appropriate exercise dosing.

## Introduction

Exercise intensity, specifically vigorous intensity, appears to function as a critical dosing ingredient for improving both peak oxygen consumption (VO₂peak)^1–3^ and walking outcomes^4–6^ after stroke. A recent systematic review and meta-analysis demonstrated high-intensity interval training (HIIT) elicits greater gains in cardiorespiratory fitness than moderate-intensity continuous training (MICT) in people with stroke.^2^ Consequently, HIIT is increasingly viewed as a promising rehabilitation strategy.

Despite this growing evidence, most prior HIIT trials share key design features that limit clinical translation. Protocols have largely relied on treadmill-based training with harness for safety^4,7^ and maximal graded exercise testing with electrocardiographic (ECG) monitoring to establish eligibility, ensure safety, and determine exercise intensity targets.^3,4,7–10^ While methodologically rigorous, this approach creates a paradox: interventions demonstrating benefit require resources unavailable in the majority of stroke rehabilitation settings and community settings.^11,12^ Maximal exercise testing is infrequently performed in routine stroke care, and reliance on it may impede adoption for safety concerns even if HIIT proves superior to MICT. Therefore, an essential next step is determining whether HIIT can be accurately prescribed using methods feasible for real-world implementation.

To address this gap, our group previously developed and validated a total-body recumbent stepper (TBRS) submaximal exercise test in healthy adults^13^ and older adults.^14^ The test demonstrates strong agreement between predicted and measured VO₂peak^15^ and high reliability^16^ in people with stroke. We have subsequently applied the TBRS submaximal test to accurately prescribe an acute bout of high-intensity interval exercise in healthy young adults, older adults, and individuals post-stroke while characterizing cerebrovascular and autonomic physiological responses.^17–21^ Submaximal testing allows individualized dosing without requiring maximal exertion, specialized equipment, or cardiopulmonary exercise testing expertise^13^ and we showed in people with chronic stroke, target heart rates reached thresholds for vigorous intensity.^19^

The Fitness After Stroke (FAST) randomized clinical trial evaluated the preliminary efficacy of short-interval, high-volume HIIT compared with MICT using a recumbent stepper modality that allows for inclusion of people with stroke who have a wide range of physical abilities.^8,22,23^ We hypothesized that HIIT would produce greater improvements in VO₂peak than MICT, with corresponding improvements in vascular measures and walking outcomes.

## METHODS

### Study Design and Oversight

The FAST trial was a single-site, randomized, parallel-group preliminary efficacy trial comparing HIIT with MICT in individuals with chronic stroke. The trial was registered at ClinicalTrials.gov (NCT05936008) before enrollment. The University of Kansas Medical Center Institutional Review Board approved all procedures, and written informed consent was obtained from all participants prior to study procedures. This randomized clinical trial is reported in accordance with the Consolidated Standards of Reporting Trials (CONSORT) reporting guideline. The full study protocol including inclusion and exclusion criteria is provided in Supplement 1 and are described in the published methods paper.^20^

### Participants

Community-dwelling adults aged 20 to 85 years with ischemic or hemorrhagic stroke ≥6 months prior to enrollment were recruited between July 2023 and December 2025 through the University of Kansas Stroke Recovery Registry, physician clinics, outpatient physical therapy clinics, and community outreach events.

### Randomization and Blinding

Participants were allocated 1:1 to HIIT or MICT using minimization^24^ stratified by lower extremity motor function, defined by the Fugl-Meyer Assessment–Lower Extremity^25^ score. The first participant was randomly assigned with subsequent participants allocated using weighted randomization with 80% probability to whichever group would improve balance. Group allocation was performed by an unblinded team member not involved in study assessments.

Outcome assessments were conducted by blinded assessors, and study statistician (RNM) was masked to group allocation. The principal investigator was blinded to primary and secondary outcomes identified in ClinicalTrials.gov. Participants were not informed of intervention labels (HIIT or MICT), specific exercise structure (interval vs continuous) or study hypotheses to minimize expectancy bias.^20^

### Exercise Interventions

Participants trained on the TBRS 3 times per week for 4 weeks with one-on-one supervision and continuous heart rate monitoring. Exercise intensity was prescribed using peak power output (PPO) derived from the TBRS submaximal exercise test. Each session included a standardized warm-up and cool-down. Rating of perceived exertion^26^ was recorded immediately after the intervention and again after cooldown. Blood pressure was taken using sphygmomanometer and stethoscope between minutes 16-17 of the intervention. Capillary blood lactate was obtained immediately post-exercise at sessions 2, 5, 8, and 11 as a marker of training intensity.^4^

The HIIT protocol consisted of repeated 1-minute high-intensity intervals alternated with 1-minute active recovery for 25 minutes. High-intensity intervals targeted 65%–95% of PPO at 90-100 steps per minute (spm) and recovery intervals were performed at 10% PPO and ∼50 spm.

The MICT protocol consisted of continuous exercise for 25 minutes at 45%–65% of PPO at 90-100 spm.

### Outcome Measures

Assessments were performed before and within one week after completion of the intervention.

### Primary Outcome

The primary outcome was change in predicted VO₂peak, assessed during the TBRS submaximal exercise test.

### Secondary Outcomes

Cerebrovascular hemodynamics were assessed using transcranial Doppler ultrasound to measure middle cerebral artery velocity (MCAv) at rest. Peripheral vascular function was assessed using brachial artery flow-mediated dilation, and arterial stiffness was measured using carotid-femoral pulse wave velocity (cfPWV).

### Tertiary Outcomes

Gait speed and walking endurance were assessed using the 10-meter walk test and 6-minute walk test, respectively.

### Intervention Acceptability

Exercise enjoyment was assessed using the 8-item Physical Activity Enjoyment Scale (PACES-8) during the post-intervention assessment visit.

### Adverse Event Monitoring

Prior to each study visit, we inquired about any medical changes that occurred since the prior visit. The safety of the intervention was evaluated by monitoring adverse events (AEs), defined as “any physical or psychological sign, symptom, or disease experienced during the study period that is temporally, but not necessarily causally, related to the intervention”.^27^ Consistent with standardized guidance for adverse event and serious adverse event reporting”,^28^ adverse events were classified using the National Cancer Institute Common Terminology Criteria for Adverse Events, version 5.0.^29^ An independent physician blinded to group assignment served as the independent adjudicator for relatedness, expectedness and severity.

### Statistical Analysis

This preliminary efficacy trial was designed to estimate effect sizes rather than test definitive hypotheses. Descriptive statistics are reported as mean (standard deviation) or median (interquartile range), as appropriate. Between-group differences in change scores were calculated with corresponding 95% confidence intervals. Effect sizes were computed as the between-group difference divided by the pooled standard deviation. Between group differences were assessed using analysis of covariance (ANCOVA) models. For the primary outcome, VO₂peak at 4 weeks was the response variable and the model was adjusted for baseline VO₂peak, group, and Fugl-Meyer score. Model fits were assessed using residual diagnostic plots and observed versus predicted plots. Analyses followed an intention-to-treat approach. Within-group changes were assessed post-hoc using paired t-tests, with p-values adjusted for multiple comparisons using the Benjamini-Hochberg procedure. Statistical analyses were performed using R Studio (versions 4.3.1 and 4.5.1).

## RESULTS

### Recruitment

Fifty-nine participants were consented, of whom 50 were eligible for randomization. One participant withdrew prior to randomization, resulting in 24 participants allocated to MICT, and 25 to HIIT (Figure 1). Participants had a mean age of 62.4 (SD=12.5, range=35-82) years. Demographic characteristics are presented in Table 1.

**Figure 1.**
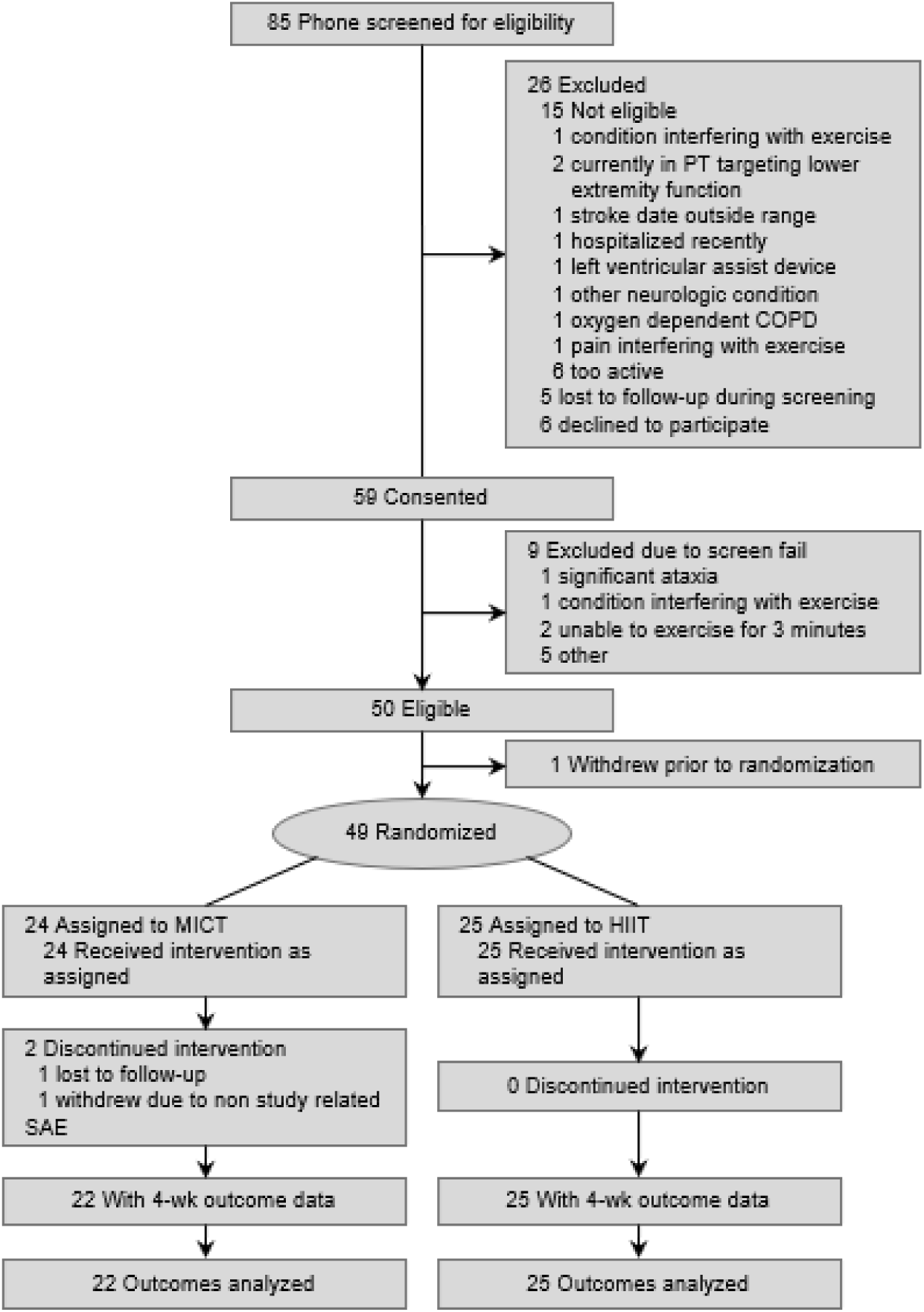
CONSORT Flow Diagram

**Table 1.** Baseline Participant Characteristics.

|  | Participants<sup>a</sup> |  |  |
| --- | --- | --- | --- |
| Characteristic | MICT (n=24) | HIIT (n=25) | p-value |
| Age, mean (SD), y | 62.42 ± 12.94 | 62.44 ± 12.33 | 0.99 |
| Sex, n(%) |  |  |  |
| Female | 12 (50) | 9 (36) | 0.32 |
| Male | 12 (50) | 16 (64) |  |
| Race, n(%) |  |  |  |
| Black/African American | 4 (17) | 7 (28) | 0.34 |
| White | 20 (83) | 18 (72) |  |
| BMI, mean (SD) | 29.13 ± 5.92 | 31.40 ± 7.97 | 0.26 |
| Education, n(%) |  |  |  |
| High school | 2 (8) | 5 (20) | 0.31 |
| Some college | 8 (33) | 6 (24) |  |
| Associate's | 1 (4) | 1 (4) |  |
| Bachelor's | 5 (21) | 9 (36) |  |
| Master's | 8 (33) | 3 (12) |  |
| Professional | 0 (0) | 1 (4) |  |
| Fugl-Meyer lower limb motor score, mean (SD)<sup>b</sup> | 29.5 ± 5.2 | 26.8 ± 5.9 | 0.10 |
| Handedness, n(%) |  |  |  |
| Right | 21 (87) | 24 (96) | 0.277 |
| Left | 3 (12) | 1 (4) |  |
| Stroke characteristics |  |  |  |
| Side of paresis, n(%) |  |  |  |
| Left | 7 (29) | 14 (56) | 0.06 |
| Right | 17 (70) | 11 (44) |  |
| Stroke Type, n(%) |  |  |  |
| Ischemic | 21 (87) | 15 (60) | 0.06 |
| Hemorrhagic | 4 (16) | 10 (40) |  |
| Stroke Location, n(%) |  |  |  |
| Left Hemisphere | 11 (46) | 7 (28) | 0.90 |
| Right Hemisphere | 6 (25) | 9 (36) |  |
| Left Subcortical | 3 (13) | 2 (8) |  |
| Right Subcortical | 1 (4) | 2 (8) |  |
| Brainstem | 2 (8) | 3 (12) |  |
| Cerebellar | 1 (4) | 1 (4) |  |
| Bilateral | 1 (4) | 1 (4) |  |
| Stroke chronicity, mean (SD), months | 30.80 ± 34.64 | 28.44 ± 24.11 | 0.78 |
| Cardiovascular Risk |  |  |  |
| Known Disease, n(%) |  |  | 0.63 |
| Cardiac disease | 2 (8) | 5 (20) |  |
| Peripheral vascular disease | 0 (0) | 1 (4) |  |
| Cerebrovascular disease | 24 (100) | 25 (100) |  |
| Type I diabetes | 0 (0) | 0 (0) |  |
| Type II diabetes | 6 (25) | 4 (16) |  |
| Renal disease | 2 (8) | 2 (8) |  |
| Smoking history |  |  | 0.52 |
| Current | 0 (0) | 1 (4) |  |
| Former | 12 (50) | 10 (40) |  |
| Never | 12 (50) | 14 (56) |  |
| CAD risk factors, n(%) |  |  | 0.67 |
| Age | 19 (79) | 24 (96) |  |
| Family History | 11 (46) | 5 (20) |  |
| Cigarette smoking | 2 (8) | 1 (4) |  |
| Physical inactivity | 22 (92) | 24 (96) |  |
| BMI | 6 (25) | 11 (44) |  |
| Blood pressure | 18 (75) | 22 (88) |  |
| Lipids | 22 (92) | 21 (84) |  |
| Blood glucose | 7 (29) | 8 (32) |  |
| HDL-C | 0 (0) | 1 (4) |  |
| Prescribed medication, n(%) |  |  | 0.27 |
| Antihypertensive | 17 (71) | 21 (84) |  |
| Statin | 20 (83) | 20 (80) |  |
| Anticoagulant | 19 (79) | 13 (52) |  |
| Antispasmodic | 4 (17) | 8 (32) |  |
| Antidepressant | 11 (46) | 8 (32) |  |
| Pain reliever | 13 (54) | 5 (20) |  |
| Stroke impact scale, mean(SD) |  |  |  |
| Raw score | 250.3 ± 20.3 | 235.4 ± 31.8 | 0.06 |
| Strength domain | 65.9 ± 20.7 | 60.2 ± 27.1 | 0.42 |
| Memory and thinking domain | 87.1 ± 7.4 | 79.9 ± 18.4 | 0.08 |
| Emotion domain | 88.5 ± 10.1 | 81.6 ± 15.1 | 0.06 |
| Communication domain | 90.9 ± 13.2 | 86.7 ± 15.5 | 0.31 |
| ADL/IADL domain | 88.3 ± 11.0 | 84.4 ± 13.1 | 0.26 |
| Mobility domain | 86.1 ± 12.8 | 83.3 ± 15.3 | 0.49 |
| Hand function domain | 71.0 ± 28.8 | 53.6 ± 38.8 | 0.08 |
| Participation/role function domain | 82.4 ± 18.3 | 74.1 ± 21 | 0.15 |

Abbreviations: BMI, body mass index; mo, month; HDL, high-density lipoprotein; CAD, coronary artery disease; ADL, activities of daily living; IADL, instrumental activities of daily living
<sup>a</sup> Data are presented as the number (percentage) of participants unless otherwise indicated
<sup>b</sup> Range, 0 to 34; higher scores indicate less motor impairment.

### Treatment Fidelity

Overall session attendance averaged 99.5% (SD=2.0%; range=91.7%–100%) with no between-group differences (HIIT: 99.0% [2.7%]; MICT: 100% [0%]; p = 0.08). In the HIIT group, participants achieved heart rate values for vigorous intensity during 80.6% (17.7%) of prescribed high-intensity intervals, reaching a mean peak heart rate of 78.8% (3.8%) of HRmax (Figure 2) and a mean postexercise lactate concentration of 3.78 (SD=1.58, range=1.4-8.6) mmol/L. In the MICT group, 67.5% (25.3%) of training time was performed within the moderate-intensity zone, with a mean heart rate of 67.3% (6.9%) of HRmax and lactate of 3.04 (SD=0.96, range=1.4-5.8) mmol/L. The lower proportion of time spent within the target HR zone in the MICT group may reflect transient decreases in HR during blood pressure measurement (Figure 2). Postexercise lactate concentration was higher in the HIIT group (mean difference=0.735, 95% CI [0.353, 1.117], p<0.001).

**Figure 2.**
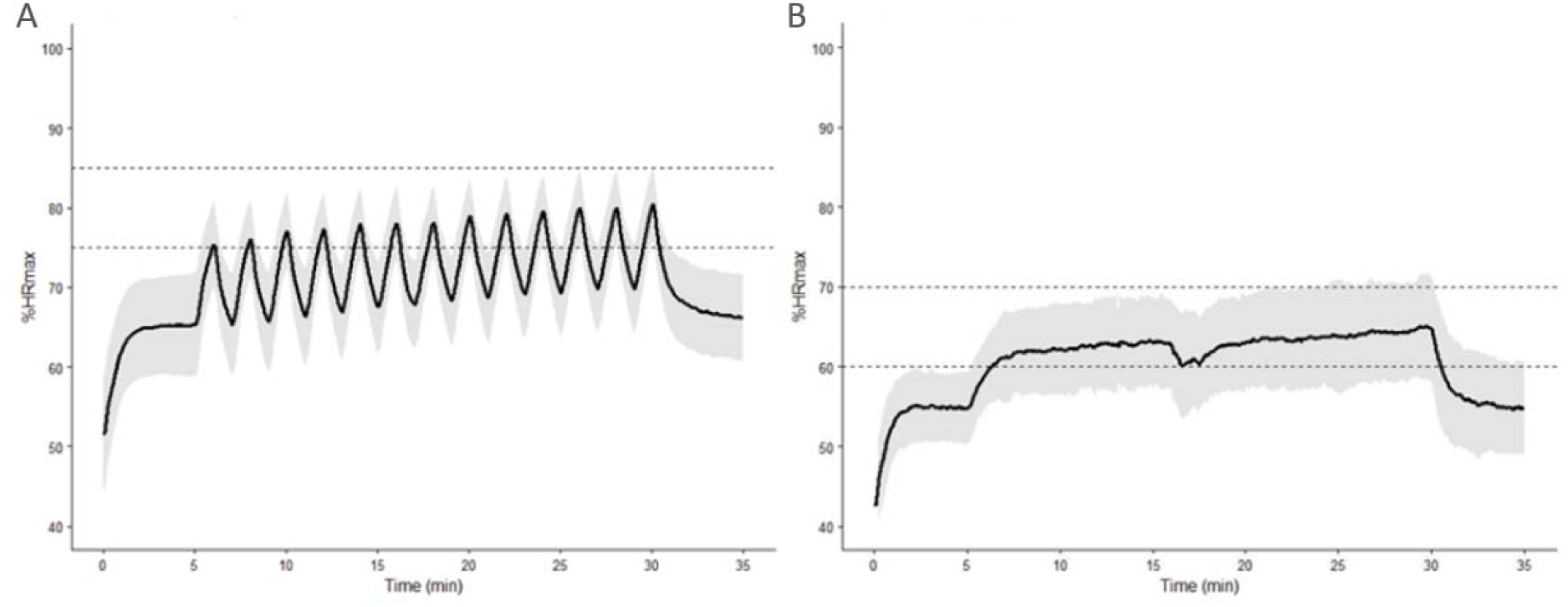
Heart Rate Responses During HIIT and MICT Sessions. Mean heart rate (black line) and SD (gray shading) across all training sessions for participants in the HIIT (A) and MICT (B) groups. Dotted lines indicate prespecified target intensity zones. %HRmax indicates percentage of age-predicted maximum heart rate. The transient decrease in heart rate in the MICT group corresponds to brief pauses for blood pressure measurement.

Exercise enjoyment did not differ between groups (PACES-8: HIIT, 47.4 [6.2]; MICT, 46.9 [7.7]; p = 0.81). However, participant comments supported high acceptability across both interventions, with descriptions of enjoyment, perceived physical benefits, improved exercise confidence, and plans to continue regular exercise after the intervention (see Supplement 1 for details). Notably, participants in the HIIT group provided more frequent and detailed feedback, particularly regarding enjoyment of the interval structure and perceived benefits of training.

### Adverse Events

Seventeen participants experienced a total of 24 adverse events (AEs), with 10 occurring in the HIIT group and 14 in the MICT group. Six AEs were classified as possibly or definitely related to the study intervention. In the HIIT group, three AEs were considered possibly related and one definitely related to the intervention. In the MICT group, two AEs were considered possibly related, one of mild severity and one of moderate severity. One serious adverse event requiring hospitalization occurred but was determined to be unrelated to the study intervention. Study-related AEs resulted in intervention modifications, including one dose reduction and three temporary pauses in the intervention. Missed visits were made up within the allotted 4-week exercise period.

### Outcome Measures

Descriptive statistics are presented in Table 2. The assumptions of the models were not violated, and overall model fit was good. There was no significant between-group effect for the primary outcome, predicted VO₂peak (β = −0.58; SE, 0.93; p = 0.54), although both groups demonstrated significant within-group improvements.

**Table 2.** Outcome Measures at Baseline and Post-Intervention.

|  | MICT |  |  |  |  | HIIT |  |  |  |  |
| --- | --- | --- | --- | --- | --- | --- | --- | --- | --- | --- |
| Outcome | n | Baseline<sup>a</sup> | Post-intervention<sup>a</sup> | Change (95% CI)<sup>b</sup> | p value | n | Baseline<sup>a</sup> | Post-intervention<sup>a</sup> | Change (95% CI)<sup>b</sup> | p-value |
| Primary outcome |  |  |  |  |  |  |  |  |  |  |
| Predicted VO<sub>2</sub> peak, mL•kg<sup>-1</sup>•min<sup>-1</sup> | 22 | 22.31 ± 8.69 | 24.34 ± 9.39 | 1.58 (0.18, 2.97) | **0.03** | 25 | 25.14 ± 11.70 | 26.27 ± 12.06 | 1.13 (0.05, 2.21) | **0.04** |
| Secondary outcomes |  |  |  |  |  |  |  |  |  |  |
| Contralesional resting MCAv, cm•sec<sup>-1</sup> | 22 | 49.01 ± 13.02 | 50.69 ± 10.79 | 1.20 (-1.95, 4.35) | 0.44 | 17 | 43.93 ± 12.47 | 44.87 ± 11.16 | -0.23 (-2.82, 2.35) | 0.85 |
| Ipsilesional resting MCAv, cm•sec<sup>-1</sup> | 19 | 48.48 ± 12.42 | 49.01 ± 12.97 | 0.54 (-1.87, 2.95) | 0.64 | 16 | 41.31 ± 12.53 | 42.42 ± 11.65 | -0.08 (-2.35, 2.18) | 0.94 |
| Stroke-affected side flow mediated dilation, % | 21 | 5.63 ± 3.31 | 6.08 ± 3.91 | 0.41 (-0.85, 1.68) | 0.50 | 23 | 4.09 ± 2.74 | 5.41 ± 3.95 | 1.43 (0.26, 2.60) | **0.02** |
| Non-affected side flow mediated dilation, % | 22 | 5.37 ± 4.13 | 6.57 ± 4.84 | 0.89 (-0.73, 2.50) | 0.27 | 23 | 4.33 ± 2.64 | 5.85 ± 3.47 | 1.60 (0.07, 3.13) | **0.04** |
| cfPWV, m•sec<sup>-1</sup> | 20 | 9.52 ± 1.88 | 9.57 ± 1.75 | -0.05 (-0.54, 0.44) | 0.83 | 22 | 9.36 ± 1.96 | 9.40 ± 2.09 | 0.06 (-0.40, 0.52) | 0.78 |
| Additional outcomes |  |  |  |  |  |  |  |  |  |  |
| Comfortable gait speed, m•sec<sup>-1</sup> | 22 | 0.96 ± 0.36 | 1.09 ± 0.36 | 0.11 (0.04, 0.18) | **0.004** | 25 | 0.90 ± 0.44 | 0.94 ± 0.39 | 0.04 (-0.02, 0.10) | 0.15 |
| Fast gait speed, m•sec<sup>-1</sup> | 22 | 1.39 ± 0.55 | 1.54 ± 0.57 | 0.11 (0.04, 0.18) | **0.004** | 25 | 1.26 ± 0.61 | 1.38 ± 0.66 | 0.12 (0.07, 0.17) | **<0.001** |
| 6-minute walk test, m | 22 | 366.25 ± 163.36 | 406.36 ± 160.28 | 28.64 (10.99, 46.28) | **0.003** | 25 | 306.00 ± 163.17 | 342.00 ± 172.12 | 36.00 (20.83, 51.17) | **<0.001** |

Abbreviations: VO<sub>2</sub>peak, peak oxygen consumption; mL•kg<sup>-1</sup>•min<sup>-1</sup>, milliliters per kilogram per minute; MCAv, middle cerebral artery blood velocity; cm•sec<sup>-1</sup>, centimeters per second; m•sec<sup>-1</sup>, meters per second; m, meters
<sup>a</sup> Baseline and post-intervention columns show the mean (SD) for each group.
<sup>b</sup> Change column shows mean difference (95% CI) from baseline to post-intervention.

No significant between-group effects were observed for resting MCAv (ipsilesional: β = −3.46; SE, 1.87; p = 0.07; contralesional: β = −0.47; SE, 1.78; p = 0.79), peripheral vascular function (flow-mediated dilation: stroke-affected arm β = 0.48; SE, 1.09; P = .66; non-affected arm β = 0.42; SE, 0.85; p = 0.62), or arterial stiffness (pulse wave velocity: β = 0.14; SE, 0.34; p = 0.68). Within-group analyses demonstrated significant improvements in flow-mediated dilation following HIIT (Table 2).

No significant between-group differences were observed for walking outcomes, including comfortable gait speed (β = −0.06; SE, 0.04; p = 0.19), fast gait speed (β = 0.03; SE, 0.04; p = 0.53), or walking endurance (β = 9.04; SE, 10.32; p = 0.39). Within-group analyses demonstrated improvements in fast gait speed and walking endurance in both groups, whereas comfortable gait speed improved only following MICT.

## DISCUSSION

This randomized clinical trial demonstrates that HIIT can be accurately prescribed and delivered after stroke without maximal cardiopulmonary exercise testing. Using a submaximal testing approach, we achieved clear separation of exercise intensity. This intensity-guided strategy enabled vigorous training with high adherence and low adverse event rates in a clinically complex cohort. Despite verified differences in training intensity and objective confirmation of workload through heart rate and lactate responses, HIIT did not produce greater improvements in predicted VO₂peak compared to MICT over 4 weeks. These findings align with the initial 4-week timepoint of the multicenter HIT-Stroke trial, in which early between-group differences were not observed and divergence emerged only with longer training durations.^4^ Collectively, these data suggest that the early aerobic response is driven by adequate dosing, whereas the incremental benefits of HIIT may require longer exposure^1,4^ and should be considered for future trials.

Prior HIIT trials have typically relied on maximal cardiopulmonary exercise testing with ECG monitoring to determine eligibility and prescribe exercise intensity. As highlighted in contemporary commentary, these requirements represent a substantial barrier to implementation in routine clinical practice.^30,31^ Further, surveys of physical therapists indicate that maximal exercise testing is rarely used in stroke rehabilitation, with only approximately 2% reporting that the majority of their patients undergo stress testing prior to exercise prescription, highlighting the limited use of maximal exercise testing in clinical practice.^11^ Therefore, by prescribing and delivering vigorous-intensity training using a submaximal testing model that does not require maximal exertion or ECG monitoring, the present study addresses this translational barrier directly. Importantly, vigorous-intensity intervals were achieved without exceeding 85% of age-predicted HRmax, demonstrating that clinically meaningful training intensity can be delivered safely using this approach. These findings support the feasibility of intensity-guided aerobic training outside specialized laboratory environments.

The magnitude of change observed in the HIIT group (+1.1 mL•kg^-1^•min^-1^) was nearly identical to that reported at the 4-week time point in the multisite treadmill-based HIT-Stroke trial (+1.3 mL•kg^-1^•min^-1^).^4^ In contrast, the improvement observed in our MICT group (+1.6 mL•kg^-1^•min^-1^) exceeded that reported in the treadmill trial at 4 weeks (+0.4 mL•kg^-1^•min^-1^), potentially reflecting higher achieved training intensity. Notably, both groups in the present study achieved improvements exceeding 1 mL•kg^-1^•min^-1^ within 4 weeks, a magnitude considered clinically meaningful for peak VO_2_.^8^ However, evidence from longer duration trials suggests that differences between HIIT and MICT may emerge over time.^4,8^ For example, Moncion et al. reported a 3.52 mL/kg/min improvement in VO₂peak following HIIT compared with 1.76 mL/kg/min following MICT at 12 weeks.^8^ These findings underscore the need for longer-duration trials to better define the optimal time course of aerobic adaptation and determine training duration required to differentiate exercise modalities after stroke.

Within the HIIT group, flow-mediated dilation improved significantly over 4 weeks, whereas no change was observed following MICT. Endothelial adaptation is a well-recognized early response to aerobic training, often preceding detectable changes in arterial stiffness or vascular structure.^32,33^ The oscillatory shear stress characteristic of interval training may provide a potent endothelial stimulus, consistent with observations in other cardiovascular populations.^34,35^ The magnitude of change observed (1.4–1.6%) is associated with reduced cardiovascular risk, underscoring the potential relevance of vigorous-intensity training for secondary prevention after stroke.^36^ However, between-group differences were not detected for other cerebrovascular or peripheral vascular outcomes. Pulse wave velocity remained unchanged, consistent with evidence that structural arterial remodeling requires longer training exposure. Resting MCAv similarly did not change, consistent with prior reports suggesting that cerebrovascular adaptations may manifest preferentially in vasomotor reactivity rather than resting velocity.^37^

Both training groups demonstrated clinically meaningful improvements in walking performance over the 4-week intervention despite not engaging in task specific treadmill training. Fast gait speed improved following HIIT and MICT. Comfortable gait speed improved following MICT, with changes meeting established minimal clinically important difference thresholds (≥0.1 m/s).^38,39^ Walking endurance also improved in both groups, with the HIIT group exceeding the minimal clinically important difference for the 6-minute walk test (≥34.4 m). Although task-specific treadmill training is often emphasized in post-stroke gait rehabilitation, the present findings support the concept of training transference, whereby repetitive lower-extremity activation during cyclic exercise translates to improved overground walking performance.^40^ Our data suggests that intensity-guided aerobic exercise using a total-body recumbent stepper can meaningfully influence walking outcomes. Importantly, the magnitude of improvement in walking endurance and fast gait speed observed over 4 weeks was comparable to, and in some cases exceeded, that reported in longer-duration treadmill-based^4^ and total-body recumbent stepper protocols,^1^ which may reflect the high level of treatment fidelity and achievement of target training intensities in the present study.

## Strengths and Limitations

This trial has several notable strengths. First, the FAST trial enrolled participants with both ischemic and hemorrhagic stroke, and women comprised 43% of the cohort, a proportion higher than that reported in many prior HIIT studies.^1,3,4,41,42^ Second, randomized allocation with concealed group assignment and blinded outcome assessment minimized risk of bias. Participants were unaware of intervention allocation, reducing expectation effects that may influence performance in trials comparing HIIT and MICT. Third, treatment fidelity was exceptionally high, with near-complete attendance and clear physiologic separation of intensity confirmed through continuous heart rate monitoring (Figure 2). Fourth, systematic adverse event collection demonstrated a low rate of intervention-related events and no study-related serious adverse events despite inclusion of a clinically complex cohort spanning a broad age range.

The intervention model further strengthens the translational relevance of the findings. By using a submaximal testing paradigm, this study operationalized vigorous-intensity training during HIIT without reliance on maximal cardiopulmonary exercise testing or ECG monitoring. Both interventions exceeded typical intensity levels previously reported in stroke rehabilitation,^43–45^ enhancing confidence that observed effects were attributable to adequately dosed aerobic stimulus rather than minimal clinical exposure. Participant-reported feedback supports the acceptability of this approach. Exercise enjoyment did not differ between groups, and qualitative responses (Supplemental data) indicated high engagement, increased exercise confidence, and intent to continue aerobic training after study completion. Several participants described improvements in physical steadiness and overall well-being, suggesting potential enhancement of exercise self-efficacy, although not directly assessed. Together, these behavioral signals strengthen the external validity and sustainability implications of intensity-guided aerobic training after stroke.

A primary limitation of the present study was the modest sample size, which may have limited our ability to detect between-group differences. As a preliminary efficacy trial, this study was designed to estimate effects and inform future trials, not definitive comparisons. Accordingly, the absence of statistically significant differences does not exclude the possibility that clinically meaningful divergence may emerge with larger sample sizes or longer intervention durations, as reported in other trials comparing HIIT and MICT. Additionally, we cannot fully exclude the potential influence of expectancy bias and that both groups exercised in their target HR zones with high adherence, which may contribute to the lack of significant findings.

Additional limitations include a single-site design and a short intervention duration of 4 weeks. The primary outcome relied on predicted rather than directly measured VO₂peak, although the submaximal testing approach was intentionally selected to enhance clinical scalability and has demonstrated validity.^13,15^ Physical activity outside supervised sessions was not objectively monitored, and long-term maintenance of training effects was not assessed. Finally, findings are limited to individuals with chronic stroke and may not generalize to earlier recovery phases.

## Conclusion

In this randomized clinical trial involving individuals with chronic stroke, vigorous-intensity HIIT delivered using a submaximal prescription model did not produce greater improvements in predicted VO₂peak than MICT over 4 weeks. However, both HIIT and MICT resulted in clinically meaningful improvements in cardiorespiratory fitness and walking performance. HIIT additionally improved endothelial function, supporting the biologic plausibility of intensity-dependent vascular adaptation. Importantly, the ability to prescribe and deliver vigorous-intensity training without maximal cardiopulmonary exercise testing supports the potential for broader integration of intensity-guided aerobic training into routine stroke rehabilitation.

## Data Availability

All data produced in the present study are available upon reasonable request to the authors.

## Author contributions

Design: SAB, EDV, RNM; Data acquisition: BLB, AE, NS, AJBC, TB, AM, ESV, EMH, EDV, SM, SAB; Analysis and interpretation: BLB, TB, RNM, EDV, MA, SM, SAB; Drafting: BLB, RNM, EDV, ESV, TB, SAB; Draft approval: BLB, AE, NS, AJBC, TB, RNM, EDV, AM, ESV, EMH, MA, SAB.

## Data access, responsibility, and analysis

SAB had full access to all the data in the study and takes responsibility for the integrity of the data and the accuracy of the data analysis.

## SUPPLEMENTARY METHODS

### Study Design

**Figure S1.**
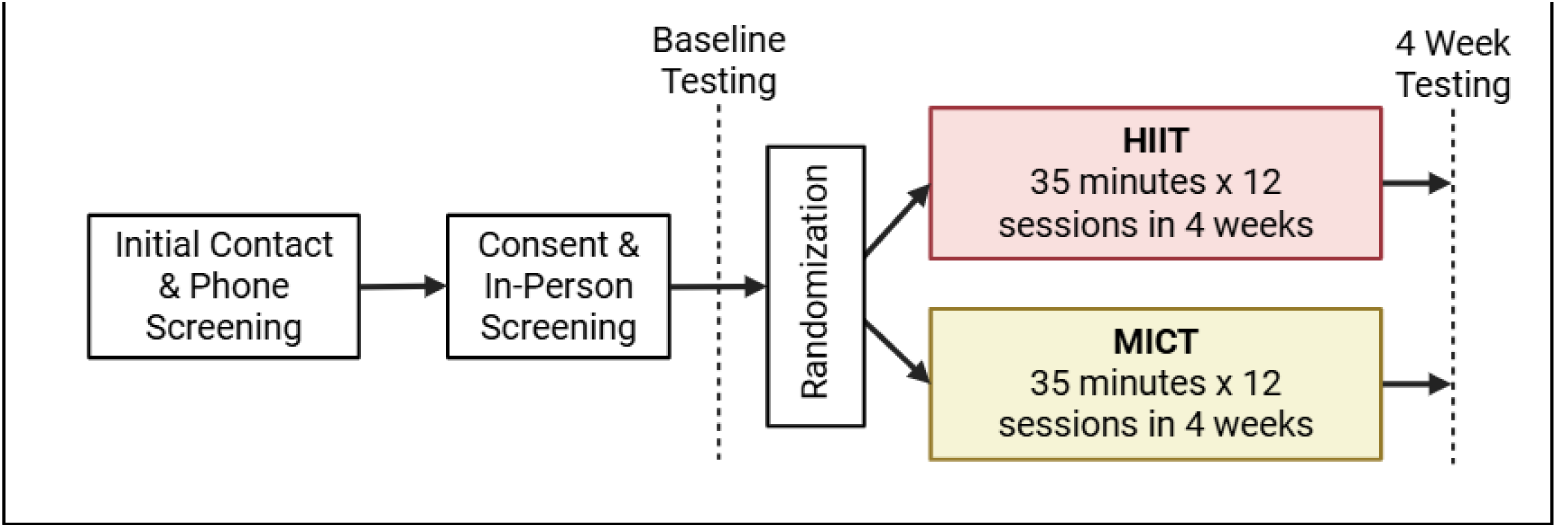
depicts the study design.

### Participants

Inclusion criteria:

- Both sexes between the age of 20-85 years at time of consent
- Chronic ischemic or hemorrhagic stroke 6 months to 15 years at consent. People with stroke and newly diagnosed cardiovascular complications had >50% prevalence of recurrent stroke at 5 years. Index stroke or recurrent stroke on same side as index stroke will be allowed.
- Walk overground with assistive devices and no continuous physical assistance from another person to perform tests for gait speed and six-minute walk test
- Exercise continuously for a minimum of 30 watts for 3 minutes on the recumbent stepper to demonstrate the ability to perform the exercise test.
- No aerobic exercise contraindications or other safety/physical concerns during the submaximal exercise test.
- Able to communicate with investigators, follow 2-step command & correctly answer consent comprehension questions
- Currently participating in less than 150 minutes of physical activity/week assessed by the Rapid Assessment of Physical Activity
- Stable blood pressure and statin medication doses for 30 days prior to enrollment due to effects on vascular health/hemodynamics

Exclusion criteria:

- Hospitalization for cardiac or pulmonary disease within past 3 months
- Implanted pacemaker or defibrillator limiting exercise performance
- Reported pain that limits or interferes with activities of daily living and physical activity/exercise
- Severe LE spasticity (Ashworth >2) due to inability to exercise
- Recent history (<3 months) of illicit drug or alcohol abuse or diagnosis of significant mental illness
- Major post-stroke depressions (Patient Health Questionnaire, PHQ-9 ≥ 10)
- Currently participating in physical therapy targeting lower extremity function or another interventional study that may influence study outcomes
- Other significant neurologic, orthopedic or peripheral vascular conditions that would limit exercise participation
- Oxygen-dependent chronic obstructive pulmonary disease
- Diagnosis of other neurological disease (Multiple Sclerosis, Alzheimer’s disease, Parkinson’s disease)
- Self-reported pregnancy

### Sample Size

The sample size of 50 was selected to provide adequate precision around the estimated between-group difference in VO_2_peak (anticipated half-width of the 95% CI ≈3.6 mL·kg⁻¹·min⁻¹, assuming a 2 mL·kg⁻¹·min⁻¹ difference and SD of 4 mL·kg⁻¹·min⁻¹).

### Randomization

Eligible participants were randomized by an unblinded study team member to high-intensity interval training (HIIT) or moderate-intensity continuous training (MICT) using minimization, stratified by lower extremity function. Lower extremity function was determined using the Fugl-Meyer Assessment, lower extremity subscale, where participants with a score ≥21 were classified as “high mobility” and those with a score <21 were classified as low mobility.^1^

### Exercise Interventions

Exercise sessions were performed with one-on-one supervision. Leg stabilizers were used as necessary for participants with lower extremity hemiparesis to promote proper leg alignment and prevent hip abduction or adduction during exercise.

During exercise, heart rate (HR) was continuously monitored using a Polar H10 chest monitor (Polar Electro Oy, Kempele, Finland). Target HR zones were calculated using age-predicted HR_max_ multiplied by target intensity. For participants not taking a beta-blocker, 220-age was used to determine age-predicted HR_max_. For individuals taking a beta-blocker, 164-(0.7*age) was used.^2^ The target intensity for MICT was 60-70% HR_max_, whereas HIIT was prescribed at 75-85% HR_max_ for HIIT. Exercise intensity was monitored using a 2-step process to optimize adherence. During exercise, HR was continuously monitored on the Polar Beat application. Following the session, post-session HR analyses were conducted in R Studio,^3^ where the exercise HR response was plotted against the target intensity zone to ensure adherence to the prescribed intensity and guide increasing exercise intensity over the 4-week training period.

Both groups exercised for 35 minutes total, with a 5-minute warm-up at 30% peak watts and a 5-minute cool-down at 20% peak watts. Blood pressure was recorded before exercise, at the mid-point of exercise, and post-exercise. Rating of perceived exertion was recorded every session immediately post-exercise using the Borg 6-20 scale.^4^ Lactate measurements were also obtained as a surrogate measure of exercise intensity at sessions 2, 5, 8, and 11 using a fingerstick and lactate meter (Lactate Plus, Nova Biomedical, Waltham, MA, USA).

### Outcome Measures

#### Primary Outcome

Peak oxygen consumption (VO_2_peak): VO_2_peak was predicted using the Total Body Recumbent Stepper (TBRS) submaximal exercise test.^5^ Participants began the test at 30 watts with a step rate of 90-100 steps per minute. As previously published,^5–8^ the watts were progressively increased until the participant reached 85% of their age-predicted HR_max_, completed all 4 stages of the test, requested to stop, or demonstrated exercise testing termination criteria.^9^ Predicted VO_2_peak was calculated using our open-access online application^10^ based on the following equation:

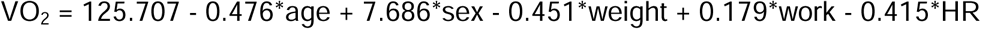

where age is in years, for sex male is 1 and female is 0, weight is in kilograms, work is the final watts used during the test, and HR is the ending heart rate during the test.

#### Secondary Outcomes

Cerebrovascular hemodynamics: Cerebrovascular hemodynamics were assessed in a dimly-lit, temperature-controlled room using transcranial doppler ultrasound (TCD; 2-MHz, Multigon Industries Inc, Yonkers, New York) at rest and during an acute bout of exercise. With the participants seated on the TBRS, we located bilateral middle cerebral artery velocity signals using a transtemporal window approach. The parameters used at baseline, including gain, depth, gate, amplitude, and probe location, were recorded and reproduced post-intervention to ensure insonation of the same middle cerebral artery branch pre- and post-intervention. Participants were also fitted with a 5-lead electrocardiogram for HR (Cardiocard, Nasiff Associates, Central Square, New York), nasal cannula for end-tidal carbon dioxide (BCI Capnocheck Sleep 9004 Smiths Medical, Dublin, Ohio), and a finger cuff for beat-to-beat blood pressure (Finapres, Medical Systems, Amsterdam, the Netherlands). The rest recording was performed using an 8-minute duration. Data were recorded using an analog-to-digital data acquisition unit (NI-USB-6212, National Instruments) and custom-written MATLAB software (R2019a or higher, The MathWorks, Inc., Natick, MA).^11–16^

Flow-mediated dilation (FMD): FMD was conducted bilaterally in accordance with current guidelines, and with 5-lead electrocardiogram gating.^17^ Participants reported to the laboratory between 7:00am and 9:00am to account for diurnal variation in vascular function. Participants were asked to fast and avoid tobacco products for ≥6 hours, avoid alcohol or caffeine for ≥12 hours, refrain from vigorous-intensity activity for ≥24 hours, and withhold blood pressure or statin medications until after FMD completion.

Following 15-minutes of supine rest, a 1-minute baseline recording of the right brachial artery was obtained. Next, FMD was performed, and a rapid inflation pneumatic cuff, 1-2cm distal to the antecubital fossa, was inflated to a suprasystolic pressure of 220 mmHg for 5-minutes. The cuff was then deflated, and the brachial artery diameter and blood velocity were simultaneously and continuously recorded for 3 minutes. Procedures were then repeated on the left arm. Baseline parameters used, including gain, depth, dynamic range, and angle of insonation (≤60°) were recorded and reproduced post-intervention. Artery diameter and blood velocity were analyzed using semi-automated edge-detection software (Brachial Analyzer, Medical Imaging Applications, Coralville, Iowa).

Pulse wave velocity: The participant remained in supine following FMD, and arterial stiffness was assessed using carotid-femoral pulse wave velocity (SphygmoCor, Itasca, IL). A pressure cuff was fitted around the participants’ upper thigh over the femoral artery. The carotid and femoral pulses were palpated by the technician, and the distances between the carotid pulse and suprasternal notch, suprasternal notch and cuff, and femoral pulse and cuff were recorded. We obtained and averaged two recordings. Pulse wave velocity was automatically calculated by our device using the following equation: pulse wave velocity (m*s^-1^) = pulse wave velocity distance/pulse transit time.

#### Additional Outcomes

Gait speed: We assessed comfortable and fast gait speed using the 10-meter walk test, where participants were instructed to walk a 14-meter path, and the middle 10-meters were timed. Participants completed two trials each at their comfortable and fast gait speeds, and the trials were averaged. Participants were instructed to use their daily orthotics and assistive devices, as applicable.

Walking endurance: We assessed walking endurance using the 6-minute walk test over a 30-meter path and aligned with the American Thoracic Society guidelines.^18^ Orthotics and assistive devices were used, as necessary.

Global and regional cerebral blood flow (CBF): Magnetic resonance imaging (MRI) with a Siemens 3T Skyra scanner was used to quantify global and regional CBF.

For CBF measurement, participant underwent on3D GRASE pseudo-continuous arterial spin labeling (pCASL) sequences. pCASL was collected with a background suppressed 3D GRASE protocol (TE/TR = 22.4/4300 ms, FOV = 300 × 300 × 120 mm^3^, matrix = 96 × 66 × 48, Post-labeling delay = 2s, 4-segmented acquisition without partial Fourier transform reconstruction, readout duration = 23.1 ms, total scan time 5:48, 2 M0 images). A T1-weighted, 3D magnetization prepared rapid gradient echo (MPRAGE) structural scan (TR/TE = 2300/2.95 ms, inversion time (TI) = 900 ms, flip angle = 9 deg, FOV = 253 × 270 mm, matrix = 240 × 256 voxels, voxel in-plane resolution = 1.05 × 1.05 mm2, slice thickness = 1.2 mm, 176 sagittal slices, in-plane acceleration factor = 2, acquisition time = 5:09) was collected for anatomical processing.

CBF was calculated using a process adapted from the Laboratory of Functional MRI Technology CBF Preprocess and Quantify packages for CBF calculation (loft-lab.org, ver. February 2019). We created individualized gray matter regions of interest (whole brain, hippocampus, and cerebellum as a reference region) for each participant using the Statistical Parametric Mapping CAT12 (neuro.uni-jena.de/cat, r1059 2016-10-28) package for anatomical segmentation (Dahnke R, Yotter RA, Gaser C. Cortical thickness and central surface estimation.^19^ We motion corrected labeled and control pCASL images separately for each sequence, realigning each image to the first peer image following M0 image acquisition. CBF was calculated with surround subtraction of averaged labeled and controlled images pair without biopolar gradients^20^ producing a subtraction image.

For global and regional CBF estimates, a whole cerebrum gray matter mask in native space was defined from the T1 MPRAGE using the CAT12 package with default parameters. The mask was coregistered to the M0 image, and coregistration parameters were applied to the Neuromorphometrics atlas associated with CAT12. Individualized global cortical and regional gray matter masks were created by convolving the gray matter probability mask ( .0.5) with the Neuromorphometrics atlas and creating individualized regions of interest. Average CBF in each subtraction volume in each mask was reported in in units of mL*100 g tissue^−1^*min^−1^. Means and range are provided in the Supplemental Table below.

### Adverse Event and Safety Monitoring

Participants were systematically assessed for adverse events (AEs) throughout the study, including both during testing and intervention. At each study contact, participants were queried about the occurrence of any untoward medical changes (AEs) regardless of perceived relatedness to the trial. Participants were asked if they had any changes in their health and then were specifically asked about any falls, injuries, pain, and fatigue. Participants were reminded to report any health changes or events occurring between scheduled contacts.

All reported AE were documented by study staff and reviewed by the study team. To promote consistency and reduce bias in classification, reported events were reviewed by an independent physician who was blinded to participant group assignment. This adjudicator evaluated event descriptions and supporting documentation to confirm event body system classification, severity grade, expectedness, and relatedness to the intervention. Discrepancies or unclear cases were resolved through review of source documentation and discussion with study investigators when necessary.

Events were categorized and graded according to the National Cancer Institute Common Terminology Criteria for Adverse Events (CTCAE), version 5.0, which provides standardized criteria for classification and severity grading.

## SUPPLEMENTARY RESULTS

### Treatment Fidelity

#### Intensity

As reported, the average peak intensity for HIIT over the duration of the intervention was 78.77(3.78)% HR_max_, and the average intensity for MICT was 67.30(6.90)% HR_max_. Table S1 shows the intensities reached by the HIIT and MICT groups across each week. Additionally, we report the rating of perceived exertion (RPE). The reported RPE directly align with vigorous- and moderate-intensity zones in HIIT and MICT, respectively.^19^

**Table S1.** Intervention Intensity by Week.

|  | HIIT |  | MICT |  |
| --- | --- | --- | --- | --- |
|  | %HRmax | RPE | %HRmax | RPE |
| **Week 1** | 78.79 (3.71) | 14.57 (2.11) | 66.63 (7.34) | 13.04 (2.27) |
| **Week 2** | 78.23 (3.95) | 14.35 (2.36) | 66.52 (7.40) | 13.16 (2.22) |
| **Week 3** | 79.20 (3.97) | 14.20 (2.61) | 67.70 (6.76) | 12.88 (2.06) |
| **Week 4** | 78.86 (3.68) | 14.31 (2.74) | 68.33 (6.35) | 12.71 (1.97) |
| **Overall** | 78.77 (3.78) | 14.36 (2.45) | 67.30 (6.90) | 12.95 (2.13) |

For HIIT, we report the average peak intensity reached during the high-intensity intervals. For MICT, we report average intensity across the 25-minutes of continuous exercise; %HRmax, percent of age-predicted heart rate maximum; RPE, rating of perceived exertion (6-20)

#### Blood Pressure

As shown in table S2, systolic and diastolic blood pressure remained in recommended limits during exercise.^20^

**Table S2.** Blood Pressure Responses.

|  | HIIT | MICT |
| --- | --- | --- |
| **Before exercise (mmHg)** |  |  |
| SBP | 121.76 (16.68) | 118.47 (18.23) |
| DBP | 73.20 (9.04) | 71.65 (9.71) |
| **During exercise (mmHg)** |  |  |
| SBP | 150.22 (18.54) | 140.25 (16.86) |
| DBP | 74.13 (8.38) | 71.77 (9.62) |
| **Post exercise (mmHg)** |  |  |
| SBP | 129.01 (17.78) | 121.28 (17.01) |
| DBP | 71.88 (8.67) | 69.40 (9.28) |

mmHg, millimeters of mercury; SBP, systolic blood pressure; DBP, diastolic blood pressure

### Magnetic Resonance Imaging

Due to an institutional equipment transition, pre- and post-MRI data was obtained for 12 participants. We report descriptive statistics, mean (range).

**Table S3.** Global and regional blood flow.

|  | HIIT (n=4) |  | MICT (n=8) |  |
| --- | --- | --- | --- | --- |
|  | Baseline | Post 4-weeks | Baseline | Post 4-weeks |
| Global cortical | 33.13 (24.80, 36.78) | 34.05 (25.98, 43.51) | 29.82 (17.63, 40.63) | 30.01 (18.62, 37.49) |
| Global cerebellar | 28.65 (22.72, 35.34) | 26.64 (17.05, 43.12) | 41.39 (16.40, 59.14) | 40.76 (24.42, 56.10) |
| M precentral gyrus |  |  |  |  |
| Ipsilesional | 29.93 (16.06, 36.19) | 29.11 (23.91, 36.84) | 27.84 (15.01, 50.20) | 23.86 (10.59, 51.05) |
| Contralesional | 31.91 (17.08, 38.64) | 31.65 (25.25, 37.28) | 26.20 (15.17, 41.25) | 23.12 (10.49, 43.91) |
| L precentral gyrus |  |  |  |  |
| Ipsilesional | 21.95 (12.62, 26.44) | 38.58 (20.91, 56.05) | 24.18 (15.66, 39.74) | 20.43 (9.43, 32.83) |
| Contralesional | 28.25 (24.31, 39.49) | 35.25 (18.50, 54.77) | 26.14 (16.72, 33.75) | 22.86 (15.86, 34.50) |
| SMA |  |  |  |  |
| Ipsilesional | 22.00 (13.46, 29.14) | 21.51 (18.98, 25.44) | 20.98 (8.77, 33.92) | 16.68 (10.00, 30.50) |
| Contralesional | 25.39 (18.32, 31.65) | 22.02 (18.51, 25.67) | 22.81 (12.15, 32.89) | 17.57 (8.24, 31.87) |
| M postcentral gyrus |  |  |  |  |
| Ipsilesional | 26.02 (18.28, 29.07) | 25.46 (19.42, 31.92) | 25.72 (12.72, 40.55) | 26.61 (16.20, 54.39) |
| Contralesional | 27.41 (15.00, 37.66) | 31.96 (23.92, 37.42) | 26.03 (14.79, 43.27) | 22.04 (8.59, 35.27) |
| L postcentral gyrus |  |  |  |  |
| Ipsilesional | 22.45 (14.88, 26.78) | 37.50 (22.40, 54.58) | 24.51 (12.36, 43.80) | 23.87 (10.24, 33.28) |
| Contralesional | 29.66 (23.43, 40.45) | 37.38 (21.59, 56.73) | 27.01 (13.31, 38.75) | 25.59 (14.20, 36.71) |
| Hippocampus |  |  |  |  |
| Ipsilesional | 38.05 (29.09, 43.83) | 25.68 (12.45, 37.55) | 37.97 (25.68, 50.38) | 36.64 (25.72, 54.23) |
| Contralesional | 35.85 (31.78, 41.57) | 31.41 (27.06, 38.04) | 33.75 (13.89, 51.03) | 35.64 (18.61, 52.85) |

M, medial; L, lateral; SMA, supplementary motor area

### Qualitative Statements

The following qualitative data were recorded by the study team members throughout the assessments and intervention and are categorized below for clarity.

#### HIIT

##### Plans to Continue Exercise

- Participant reported that he plans to continue exercising upon study completion.
- Participant reported that she enjoyed the exercise sessions and is, “planning on establishing an exercise routine.”
- Participant joined a community center with a gym to continue exercising after intervention cessation, as he felt that his “walking is better,” and he “overall feels better.”
- Participant reported feeling, “bittersweet about the exercise sessions ending.” She said she was, “proud of the work” she did and plans to continue exercising after study completion.
- Participant reported that she wants to continue exercising once the study is finished.
- Participant stated, “I find this exercise pleasurable. I can’t wait to do it at home… It’s very gratifying. In fact, I am looking forward to doing it more because of the benefits I am seeing. You guys [study team] have enriched my life both mentally and physically.”

##### Enjoyed Interval-Style of Exercise

- Participant reported enjoying the interval-style exercise.

- Participant reported that she was enjoying the “up/down style of exercise.”

- Participant reported liking the intervals, as they made the exercise “more exciting.” He also reported that the active recovery intervals “were very helpful.”

##### Physical Benefit

- Participant stated, “After the exercise, I do feel steadier.”
- Participant stated, “That stepper really helps my legs!”

##### Enjoyment

- Participant reported that she was looking forward to her next session.
- Participant reported that she was sad that the session was her last, because she had “enjoyed it so much.”
- Participant reported that he “really enjoyed the study,” and is hopeful that he, “can participate in future studies.”
- Participant stated that he wished the intervention was longer, because he “enjoyed it so much.”
- Participant reported enjoying the exercise sessions.
- Participant reported that he wishes he could start the exercise intervention all over again once he finished.

##### Additional Comments

- Participant stated, “This gives me a reason to get up in the morning.”
- Participant reported feeling fatigued after an exercise session, but stated it was a “good tired.”

#### MICT

##### Plans to Continue Exercise

- Participant plans to continue exercise after study completion, with the goal of joining a gym and “doing aerobic exercise three times per week.”

##### Enjoyment and Confidence

- Participant reported enjoying the exercise sessions, and that the sessions are increasing her exercise confidence. She reported jogging “approximately 1 mile from the park” to her home with her kids, because she now “felt safe to do so.”
- Participant stated, “This has been a really good experience. I feel like I can exercise now and not be afraid to sweat.”

##### Additional Comments

- Participant reported that they feel like the exercise “might be getting easier.”

